# Evaluation of the depth and width of normal fetal Sylvian fissure by trans-cerebellar section

**DOI:** 10.1101/2021.05.07.21256819

**Authors:** Xu Pingping, Zhang Dirong, Shi Yu, Kong Fengbei, Yao Chunxiao, She Ying, Wu Guoru

**Affiliations:** Ultrasound Department, Peking University Shenzhen Hospital, Shenzhen, Guangdong, China

**Keywords:** Fetus, Sylvian fissure, depth, width, trans-cerebellar section

## Abstract

**Objective:** On the basis of retrospectively analysis the trans-cerebellar section showing the Sylvian fissure of normal fetus is better than trans-thalamic section in middle and late trimester, we prospectively established the normal reference range of developmental parameters related to the Sylvian fissure of normal fetus in the trans-cerebellar section in order to provide valuable information for the diagnosis of fetal cerebral cortical dysplasia.

**Methods:** A prospective cross-sectional study was conducted on 845 normal fetuses at 21 to 32 weeks of gestation from January 2019 to September 2020. The depth and width of the Sylvian fissure was measured respectively, and regression analysis was performed according to different gestational age groups.

**Results:** The depth of Sylvian fissure was positively correlated with gestational weeks, and the width of Sylvian fissure was negatively correlated with gestational weeks, with correlation coefficients 0.751 and 0.825 respectively (all P < 0.001).Taking gestational weeks as the independent variable, and the depth and width of Sylvian fissure of fetus as the dependent variables, linear regression analysis showed that there was a linear relationship between the independent variables and the dependent variables. The obtained equation was simplified to a formula: Fetal Sylvian fissure depth (mm)=7+0.6× (gestational weeks-21), Sylvian fissure width (mm)=8-0.6× (gestational weeks-21).

**Conclusion:** The reference range of the depth and width of Sylvian fissure of normal fetus at 21 to 32 weeks of pregnancy through the trans-cerebellar section was established, which could provide valuable information for the evaluation of the normal development of the lateral fissure of fetus and the prenatal diagnosis of fetal cortical hypoplasia.

## INTRODUCTION

Cerebral cortical developmental malformation is difficult to diagnose by prenatal ultrasound^1^. However, it seriously affects the quality life of children and could lead to severe intellectual retardation and intractable epilepsy^2^. Deformity of fetal cerebral cortex is usually manifested as abnormal sulcus development, which can be specifically manifested as delayed sulcus development, non-development of sulcus and overdevelopment of sulcus^3^. The delay of sulci development means that sulci development is more than 3 weeks later than that of normal healthy fetus of the same age^4^. The Sylvian fissure, located on the lateral side of the cerebral hemisphere, is one of the main cerebral sulcus. The Sylvian fissure is the earliest one appeared in the development of all the sulci^5^, and its development is representative. Therefore, the development law of the Sylvian fissure can be further indicated by studying the development law of the cerebral cortex. Previous studies have reported the parameters related to the Sylvian fissure in normal fetuses, but most of these studies selected the trans-thalamic section to evaluate the Sylvian fissure, and the measurement sites and methods of the relevant parameters were not uniform^6-10^. In our previous retrospective study, we have found that the Sylvian fissure was shown clearer on trans-cerebellar section than trans-thalamic section. Therefore, the aim of this study was to establish the normal reference range of the depth and width of Sylvian fissure of normal fetuses in middle and late trimester by the trans-cerebellar section, so as to provide valuable information for the evaluation of the normal development of fetal Sylvian fissure and the suspicious diagnosis of fetal cerebral cortical dysplasia by prenatal ultrasound.

## METHODS

This was a prospective cross-sectional study. Normal fetuses undergoing routine prenatal ultrasonography in our hospital from January 2019 to September 2020 were selected as subjects. To exclude abnormal fetuses and ensure the relative accuracy of gestational age, the inclusion criteria were as follows: the date of the last menstruation was clear or the gestational week was corrected by crown-lump length;21 to 32 weeks of gestation; low risk pregnancy including pregnant women without diabetes, hypertension and other pregnancy risk factors. Exclusion criteria were as follows: fetuses with known malformations or chromosomal abnormalities; the large difference between the biological gestational age and the actual gestational age of the fetuses; multiple pregnancies; nervous system abnormality occurs within 6 months after delivery.

This study was approved by the Peking University Shenzhen Hospital and Health Service Human Research Ethics Committee. All pregnant women gave informed consent to this study. The pregnant women ranged in age from 20 to 46 years old, with an average age of 27 years old. The gestational age ranged from 21 to 32 weeks, with an average gestational age of 27 weeks.

The ultrasound machine used in this study was a Voluson E8 or E10 (GE Healthcare Ultrasound) with a 4-8MHz transabdominal 2D transducer. In each examination, the biparietal diameter, head circumference, abdominal circumference, femur and other parameters of the fetus were measured to comprehensively evaluate the biological gestational age of the fetus, and then the screening of the structural malformation of each fetal system was conducted.

When detailed screening of fetal central nervous system structures, the depth and width of Sylvian fissure of the fetus were measured by trans-cerebellar section. Standard trans-cerebellar section^11^: the probe clearly showed the midline of the brain, cavity of septum pellucidum, thalamus, bilateral cerebellar hemispheres and the vermis of the cerebellum, posterior fossa and Sylvian fissure. The measurement method was shown in Figure 1. Sylvian fissure depth: made a vertical line perpendicular to the top edge through the midpoint of the top edge of the Sylvian fissure, and measured the distance from the top edge to the medial surface of the skull on the vertical line. Sylvian fissure width: made a line parallel to the top edge through the middle point of the depth line of the Sylvian fissure and measured the distance between the front and back medial edges of Sylvian fissure on the parallel line. Each patient was measured by one experienced sonographer, and each was measured 3 times. When the fetus had poor image acquisition due to positional limitations, the pregnant woman was instructed to perform the scan after moderate activity.

**Fig. 1.**
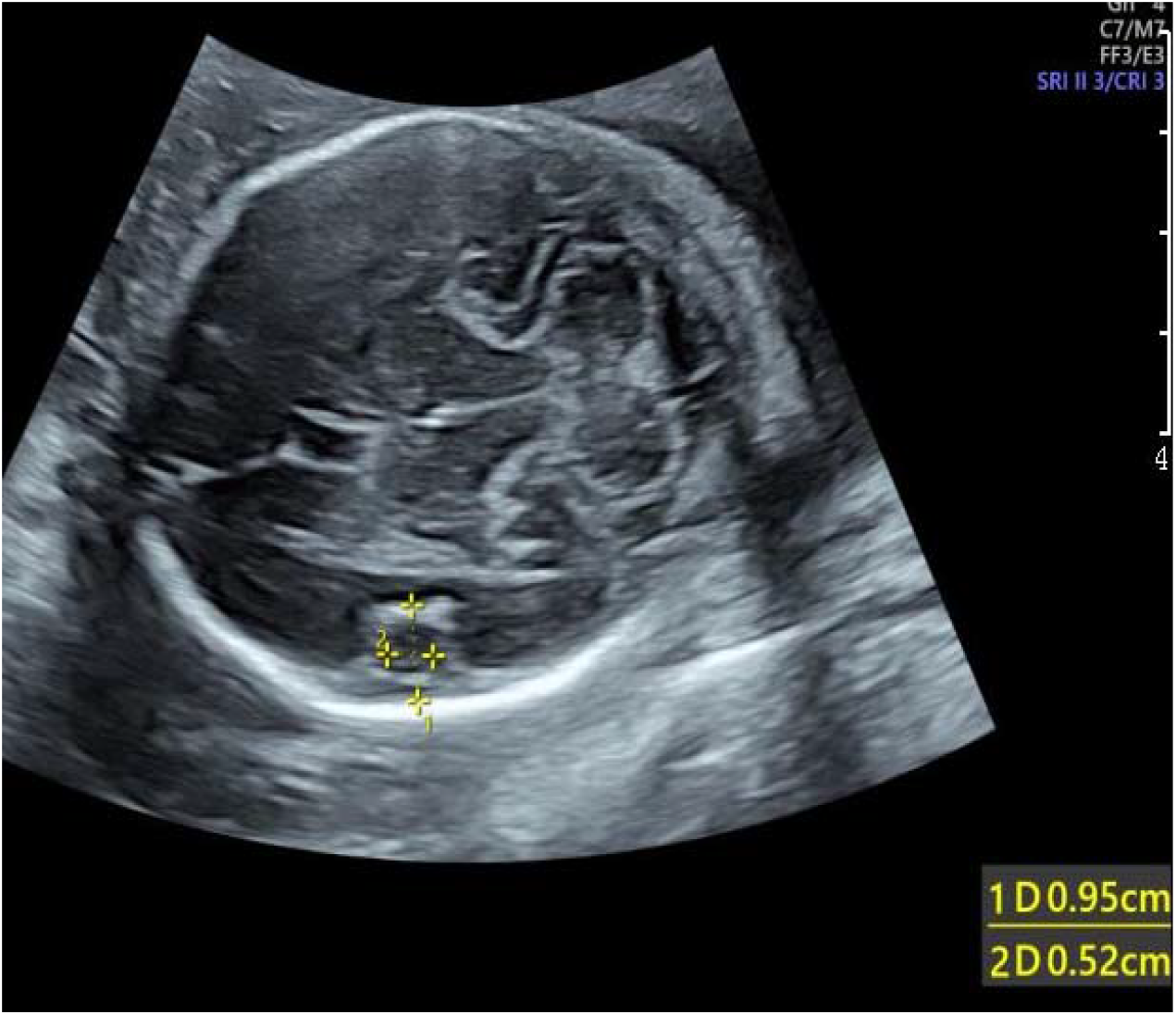
Measurement of depth and width of Sylvian fissure at 24 weeks of gestation. 1: Sylvian fissure depth; 2: Sylvian fissure width

SPSS 26.0 statistical software was used to analyze and process the data. The main steps were as follows:(i) All measured data were counted and divided into 12 groups according to gestational age. The gestational age sample size of each group was counted, the mean value and standard deviation of each parameter in each group 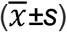 were calculated;(ii)the normality of each parameter value was tested by the Anderson-Darling test; (iii) for data that conformed to a normal distribution, the 95% reference range was calculated using the two-sided boundary value normal distribution method; for data with a skewed distribution, the normal reference range was calculated using the two-sided boundary value percentile method. The scatter plot of the relationship between the depth and width of Sylvian fissure of fetus and the week of gestation was drawn, and a linear regression equation was established by linear regression analysis, with *p* < 0.05 being statistically significant.

## RESULTS

A total of 845 subjects were included in the study. The sample data of the depth and width of Sylvian fissure of fetus at different gestational weeks were consistent with or approximately normal distribution. The mean value, standard deviation and 95% reference range of each parameter were calculated at different gestational weeks, and the bilateral boundary value was taken. The data were shown in Table 1 and Table 2.

**Table 1.** The mean value, standard deviation 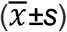 and 95% reference range of the depth of Sylvian fissure of fetus at different gestational weeks

| Gestational weeks | Numbers | Sylvian fissure depth(mm) |  |
| --- | --- | --- | --- |
| | | ($\bar{x} \pm s$) | 95% reference range |
| 21 | 65 | 6.76±1.09 | 5.01~8.80 |
| 22 | 70 | 7.60±1.25 | 5.23~9.47 |
| 23 | 91 | 8.49±1.56 | 5.80~11.34 |
| 24 | 75 | 8.81±1.59 | 5.50~11.67 |
| 25 | 68 | 9.19±1.75 | 5.84~12.48 |
| 26 | 78 | 10.58±1.27 | 8.21~12.85 |
| 27 | 75 | 10.95±1.63 | 8.40~14.00 |
| 28 | 70 | 11.21±1.55 | 8.80~14.43 |
| 29 | 71 | 12.07±2.02 | 7.14~14.88 |
| 30 | 67 | 12.43±2.12 | 7.95~16.05 |
| 31 | 60 | 12.50±2.11 | 8.12~16.23 |
| 32 | 55 | 13.10±1.94 | 9.39~16.54 |

**Table 2.** The mean value, standard deviation 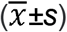 and 95% reference range of the width of Sylvian fissure of fetus at different gestational weeks

| Gestational weeks | Numbers | Sylvian fissure width(mm) |  |
| --- | --- | --- | --- |
| | | ($\bar{x} \pm s$) | 95% reference range |
| 21 | 65 | 8.31±1.27 | 6.38~10.33 |
| 22 | 70 | 7.11±1.26 | 4.93~8.70 |
| 23 | 91 | 6.88±1.45 | 4.44~9.16 |
| 24 | 75 | 6.38±1.48 | 4.00~8.64 |
| 25 | 68 | 6.23±1.51 | 3.44~8.36 |
| 26 | 78 | 5.48±1.32 | 3.07~7.67 |
| 27 | 75 | 5.07±1.56 | 2.47~7.22 |
| 28 | 70 | 3.66±1.25 | 1.80~5.54 |
| 29 | 71 | 3.02±1.19 | 1.70~5.01 |
| 30 | 67 | 2.63±1.23 | 1.10~5.34 |
| 31 | 60 | 1.88±1.01 | 0.22~3.83 |
| 32 | 55 | 1.50±1.04 | 0.15~3.65 |

Scatterplots of the relationship between the depth and width of the Sylvian fissure of the fetus and gestational weeks were drawn (Fig.2, Fig.3). Correlation analysis showed that the depth of Sylvian fissure was positively correlated with gestational weeks, with a correlation coefficient 0.751 (P < 0.001); the width of Sylvian fissure was negatively correlated with gestational weeks with a correlation coefficient 0.825 (P < 0.001).

**Fig. 2.**
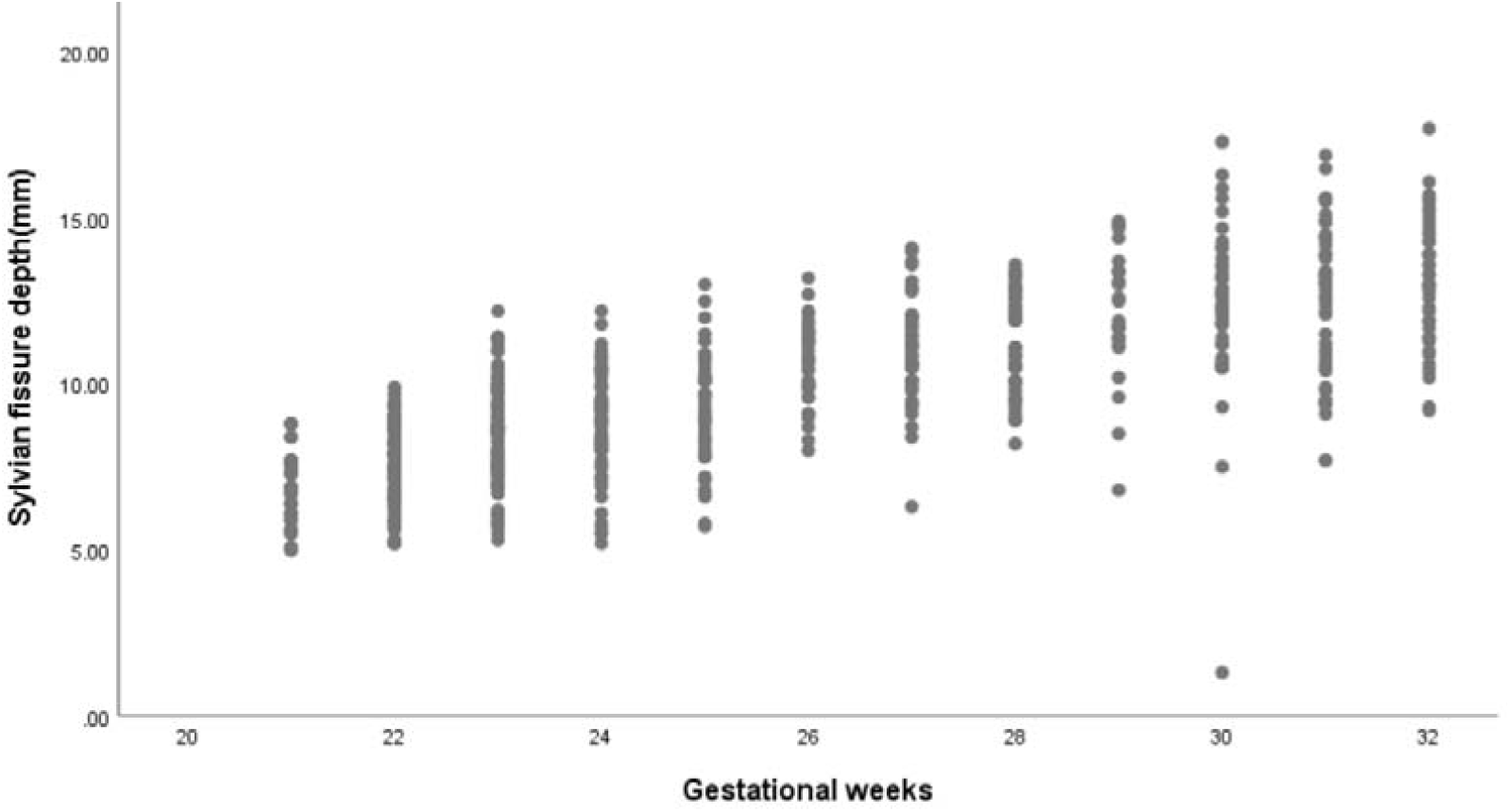
Scatterplot of the relationship between the Sylvian fissure depth and gestational weeks

**Fig. 3.**
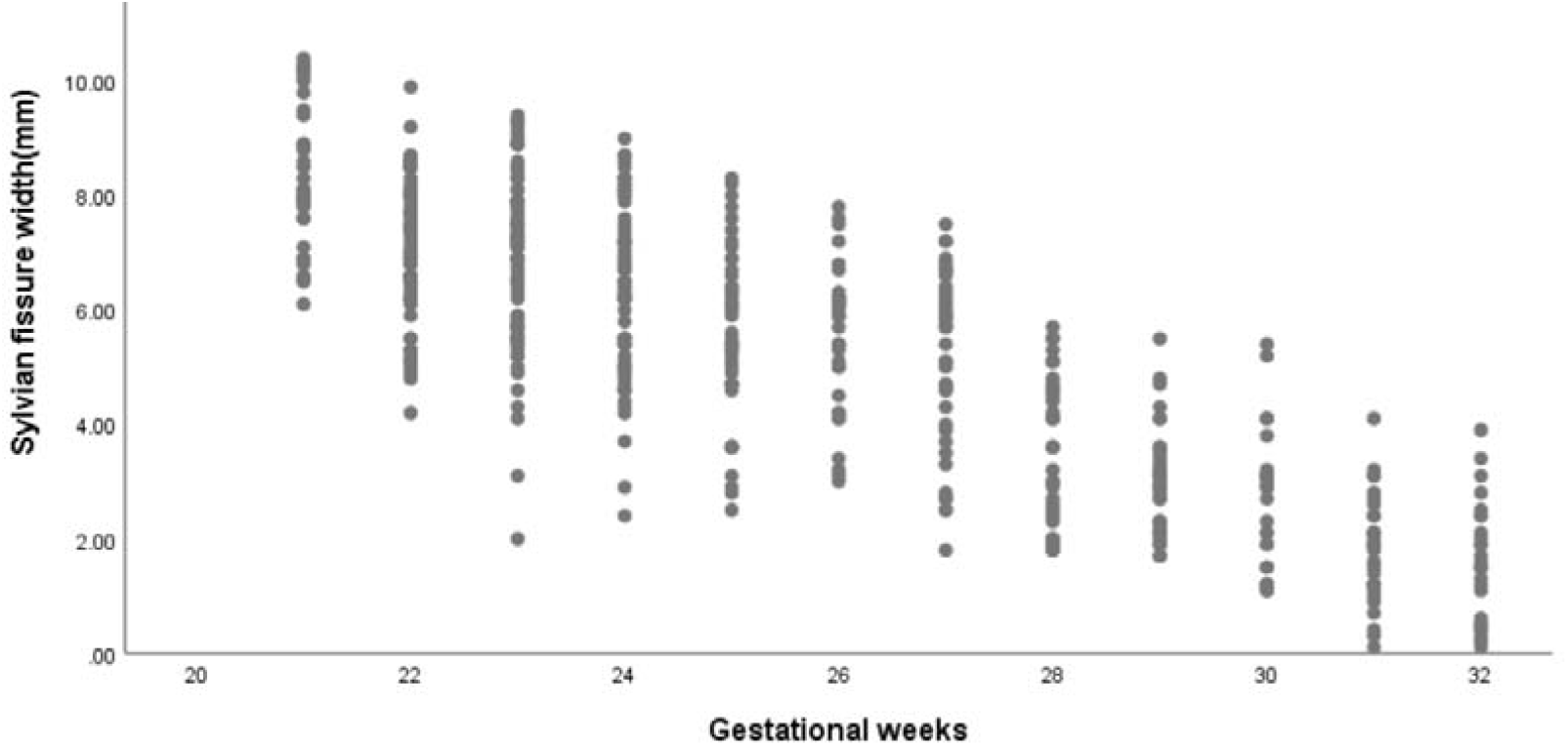
Scatterplot of the relationship between the Sylvian fissure width and gestational weeks

Taking gestational weeks as the independent variable, the depth and width of Sylvian fissure of fetus were used as the dependent variables for linear regression analysis. The results showed that there was a linear regression relationship between gestational weeks and the depth and width of the Sylvian fissure of fetus. The obtained linear regression equations were as follows: Sylvian fissure depth(mm)=7.174+0.568×(gestational weeks-21), Sylvian fissure width(mm)=8.089-0.602×(gestational weeks-21).The above equations were simplified as follows: Sylvian fissure depth(mm)=7+0.6×(gestational weeks-21), Sylvian fissure width(mm)=8-0.6×(gestational weeks-21).

## DISCUSSION

Human brain development is extremely complex, but it follows strict developmental rules. Some scholars consider that cerebral sulci can be used as a sign of fetal cranial development^12^. The developmental process of cerebral sulci can be used as a standard to evaluate the gestational age. By observing the development of cerebral sulci to further evaluate the development of the whole fetal brain is of great significance to the evaluation of fetal brain development and the diagnosis of diseases. The brain develops rapidly and has obvious morphological changes. In appearance, it can be seen that the brain develops from a tubular structure to a surface covered with furrows, and various branches of the furrows will appear in the process of forming sulci^13-15^.

The Sylvian fissure, located on the lateral side of the cerebral hemisphere, is one of the major cerebral sulcus and the earliest one sulci to appear. It separates the upper frontal and parietal lobes from the lower temporal lobes. The development of the Sylvian fissure also indirectly reflects the development speed of the frontal, parietal and temporal lobes^16^. Prenatal ultrasound can clearly show the Sylvian fissure, so it can be used to evaluate the development of the Sylvian fissure. At present, there have been reports on the evaluation of fetal Sylvian fissure, but the research conclusions are not uniform because of the differences in the study section and measurement methods. the existing studies to evaluate the development parameters of fetal fissure lateral mainly select the cross-section of the thalamus. Current studies have assessed the developmental parameters of the Sylvian fissure of fetus mainly by trans-thalamic section. However, in our previous retrospective study, we have found that the trans-cerebellar transverse section showed the Sylvian fissure clearer than the trans-thalamic section in each gestational week, which indicated that most of the developmental parameters related to the Sylvian fissure can be measured in the trans-cerebellar section clearly, and the measurement results will be more repeatable. The overall clarity of the Sylvian fissure shown on the trans-thalamic section is low, which indicated that in most cases, adjustments are needed on this standard section to measure the parameters related to the Sylvian fissure, which may affect the repeatability of the measurement results.

Therefore, in this study, the developmental parameters related to the Sylvian fissure of fetus were quantitatively measured by trans-cerebellar section and the measurement method of the depth and width of the Sylvian fissure was clearly defined.

In this study, normal fetuses from 21 to 32 weeks of gestation were selected as the research object, and trans-cerebellar section was selected and the data were successfully measured with a rate of 95.6%. The results of this study showed that the depth of Sylvian fissure was positively correlated with gestational weeks, and the width of Sylvian fissure was negatively correlated with gestational weeks, with correlation coefficients 0.751 and 0.825 respectively (all P < 0.001). Taking gestational weeks as the independent variable, and the depth and width of Sylvian fissure of fetus as the dependent variables, linear regression analysis showed that there was a linear relationship between the independent variables and the dependent variables. The obtained linear regression equations were as follows: Sylvian fissure depth(mm)=7.174+0.568×(gestational weeks-21),Sylvian fissure width(mm)=8.089-0.602×(gestational weeks-21).The above equations were simplified as follows: Sylvian fissure depth(mm)=7+0.6×(gestational weeks-21), Sylvian fissure width(mm)=8-0.6×(gestational weeks-21).During prenatal ultrasound examination, if the depth or width of the Sylvian fissure of the fetus is significantly lower than that of gestational week (depth < the 5th percentile, width > the 95th percentile), it is necessary to be alert to the possibility of fetal sulcus development retardation (as shown in Fig.4). MRI examination of the brain of the fetus confirmed cortical dysplasia.

**Fig. 4.**
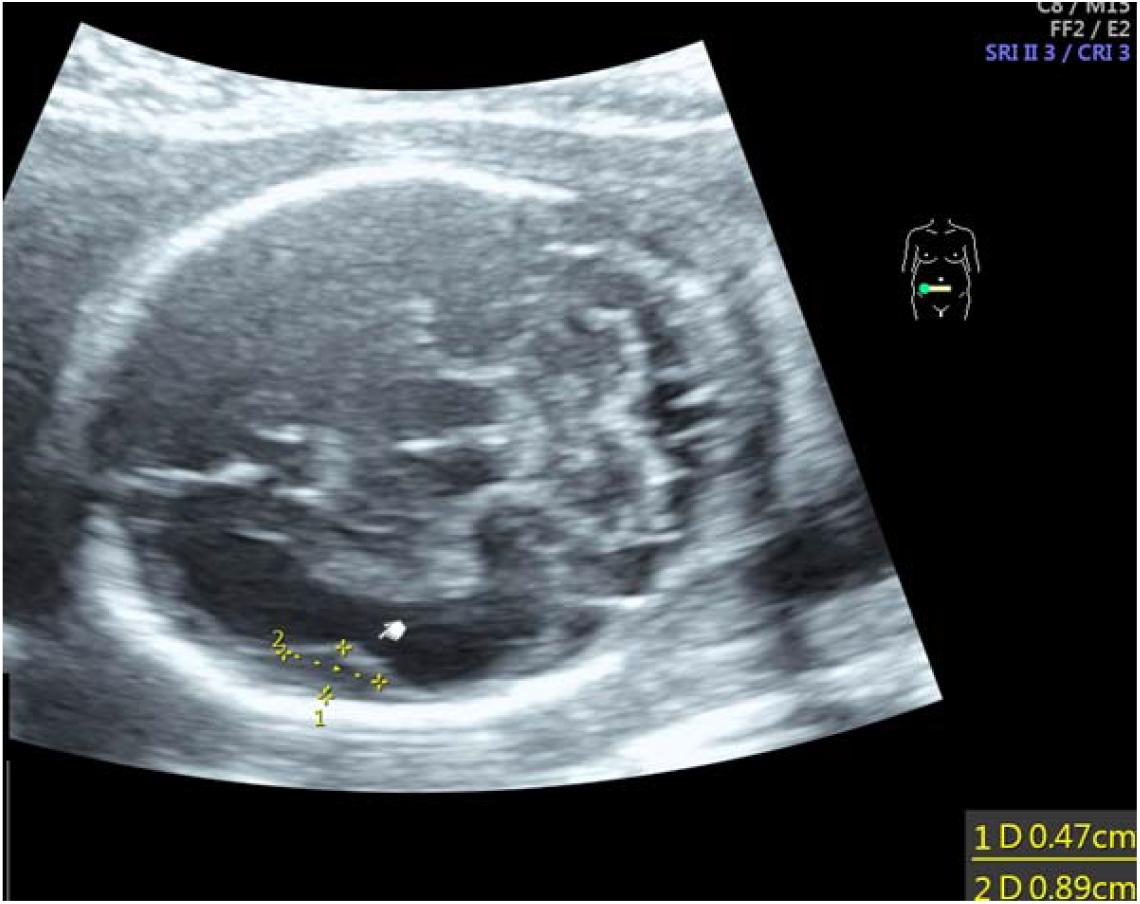
The depth of Sylvian fissure was below the 5th percentile and its width was above the 95th percentile at 24 weeks of gestation.

In this study, we innovatively selected trans-cerebellar section to evaluate the Sylvian fissure of normal fetuses and preliminarily elucidated the depth and width of the Sylvian fissure of normal fetuses at 21 to 32 weeks of gestation, which can provide valuable information for the evaluation of the normal development of fetal Sylvian fissure and the prenatal diagnosis of fetal cortical dysplasia.

## Data Availability

Data availability. The full data that supports the findings of this study are available from the corresponding author upon reasonable request.

## ACKNOWLEDGEMENTS

The authors thank all the women who participated in the study and acknowledge their significant contribution. Thank you to Zhang Dirong and Shi Yu for helping to design the research methods, Kong Fengbei and Yao Chunxiao for support with statistical analysis, She Ying and Wu Guoru for proposing some strategies for writing.

